# Impact of reducing childhood poverty on social vulnerability in young adulthood: a simulation study based on nationwide life-course data

**DOI:** 10.1101/2025.07.02.25330720

**Authors:** Signe K. Bennetsen, Leonie K. Elsenburg, Theis Lange, Adrian G. Zucco, Tjeerd Rudmer de Vries, Naja Hulvej Rod

## Abstract

**Background:** UN Sustainable Development Goal 1.2 targets a 50% reduction in childhood poverty by 2030. This study explores the potential impact of such a reduction in Denmark on (1) the distribution of individuals over trajectories of childhood adversity and (2) social vulnerability (mental health, crime and social benefits) in young adulthood.

**Methods:** We included 607,308 individuals born in Denmark (1987-1996). Childhood poverty was defined as an annual equivalized disposable income below 50% of the national median. Using group-based multi-trajectory modeling, five childhood adversity groups were previously identified based on 12 adversities (including poverty) over three dimensions from ages 0–15. A 50% annual poverty reduction was simulated, and individuals were reassigned to adversity groups. Social vulnerability (ages 16-24) was measured using three binary indicators: mental health problems, crime conviction, and long-term social benefit use. Using G-computation, we estimated how the reassignment to adversity groups affected each outcome.

**Findings:** Of those affected by the simulated poverty reduction, 32% changed adversity group. This resulted in a modest decrease in social vulnerability among these individuals: 11.6 fewer crime convictions (CI95%: 10.9–12.2), 12.6 fewer long-term benefit recipients (CI95%: 12.0–13.2), and 7.0 fewer mental health cases (CI95%: 6.2–7.8) per 1,000 individuals. Population-wide risk differences were smaller (1–2 cases per 1,000 individuals).

**Interpretation:** Reducing childhood poverty may lower social vulnerability in young adulthood, but it needs to be integrated with broader, multifaceted approaches that support children facing multiple, clustered adversities while growing up and as they transition to adulthood.

**Funding:** European Research Council (ERC) (Grant agreement No. 101124807). The Copenhagen Health Complexity center is funded by TrygFonden.

## Introduction

In 2015, UN member states committed to UN Sustainable Development Goal (SDG) 1.2 targeting a 50% reduction in poverty, including childhood poverty, by 2030 (1). Even in high-income countries, childhood poverty remains common, with about one in eight children in OECD member states living in relative poverty (2).

Children growing up in poverty are at elevated risk of developing health problems and experiencing social challenges while transitioning into adult life (3,4). This pattern of inequality emerges from a complex interplay of social and health-related factors accumulating in families experiencing poverty. For example, studies have demonstrated that children living in poverty are more likely to experience other adversities such as abuse or neglect (5), parental alcohol or drug abuse (6), severe illness or death in the family, and parental separation – experiences that may shape psychosocial development and influence future life prospects (7).

Children facing adversities have higher odds of being high-intensity users of the healthcare system, the social security system, and the justice system (8). This pattern is especially pronounced among children exposed to high and increasing adversities during childhood. These findings highlight the individual and societal costs associated with early adversity and suggest that childhood adversity creates vulnerability in the transition into adulthood.

Poverty has been suggested to exacerbate the detrimental effects of other childhood adversities, and reducing poverty may serve as a leverage point to mitigate the long-term social and health consequences associated with early adversity (9). Experimental and quasi-experimental studies have emphasized that income increments for low-income families can positively influence child development, health, and educational attainment (10). However, evidence from such studies on the longer-term effects of reducing poverty remains limited (11).

To address this gap in the literature, we investigate the impact of hypothetically reducing annual childhood poverty in Denmark by 50% per year in alignment with SDG goal 1.2. Using nationwide register data, we explore how such reductions in poverty affect the distribution of individuals over common trajectories of childhood adversity in the Danish population. Additionally, we utilize causal inference methods to simulate the influence of these reassignments following the poverty reduction on three key indicators of social vulnerability in young adulthood: long-term social benefit use, mental illness, and crime. We hypothesize that reducing childhood poverty will have long-term implications for social vulnerability in the young adult population.

## Methods

### Study population

This population-based cohort study is based on data from the Danish life course (DANLIFE) cohort, which covers 2,223,927 individuals born in Denmark between Jan 1, 1980, and Dec 31, 2015, and includes comprehensive data on childhood adversities, and a broad variety of indicators of health and socioeconomic position. The current study included all individuals born in Denmark from January 1, 1987, to December 31, 1996, excluding those who emigrated (n=30,845) or died before age 16 (n=5,906). We further excluded 3,759 individuals due to missing information on childhood poverty or covariates. This resulted in a final study population of 607,308 individuals (see flow chart in the appendix p 2).

Access to pseudonymized register data was given by the Danish Health Data Authorities and Statistics Denmark. The unique Civil Personal Registration (CPR) number, assigned to every Danish citizen at birth, enabled linkage of data on children and their parents and siblings across registries. The Danish Data Protection Agency has authorized the DANLIFE study through a shared notification issued to The Faculty of Health and Medical Sciences at the University of Copenhagen (record number 514-0641/21-3000).

### Childhood poverty

Information on poverty at every age from 0-15 years was retrieved from the Income Statistics Register (12). As extreme poverty is rare in high-income countries, we investigated *relative childhood poverty*, defined as living in a family with significantly fewer economic resources than the societal norm. Relative childhood poverty (hereafter referred to as *childhood poverty*) was operationalized as living in a family with an equivalized disposable income below 50% of the national income median in a year. This income metric is adjusted for the number of individuals living in a family (13), and the definition aligns with the OECD definition of relative income poverty (14).

### Childhood adversity trajectories

Five groups of childhood adversity trajectories were identified previously using group-based multi-trajectory modelling (15). This approach enables identification of groups of individuals with similar trajectories over time. The clustering of individuals was based on their annual rates of exposure to 12 childhood adversities from birth to age 15 across three predefined dimensions: material deprivation (i.e., childhood poverty, parental long-term unemployment), loss and threat of loss (i.e., parental somatic illness, sibling somatic illness, death of a parent, death of a sibling), and family dynamics (i.e., foster care replacement, parental or sibling psychiatric illness, parental substance or alcohol abuse, parental separation).

The group-based multi-trajectory modeling process yielded posterior probabilities for each individual, indicating their likelihood of belonging to each of the five childhood adversity groups. Each individual was assigned to the group they had the highest likelihood of belonging to. The childhood adversity groups included: Low adversity, (i.e., low rates of adversity across all three dimensions, 53%), Early material deprivation (i.e., high rates of material deprivation until age 4-5 years, 23%), Persistent material deprivation (high rates of material deprivation throughout childhood, 12%), Loss or threat of loss (i.e., high and increasing rates of adversities within the loss and threat of loss dimension, 9%), and High adversity (i.e., high and increasing rates of adversity across all three dimensions, 3%). Further details about the childhood adversity groups and the group-based trajectory modelling process are provided in the appendix (pp 3-6).

### Indicators of social vulnerability in young adulthood

We used three binary indicators of social vulnerability from 16 to 24 years: long-term social benefit use, mental health problems, and crime conviction. Every individual was followed up until their 25^th^ birthday, death or emigration.

Use of social benefits was based on information from the Danish Register for Evaluation of Marginalisation (DREAM) which includes information on weekly use of social benefits (16). We included all social benefits provided to individuals outside the workforce due to health conditions or unemployment. Long-term social benefit use was defined as receiving social benefits for 52 consecutive weeks or more during follow-up.

Mental health problems were defined as diagnosis from public and private hospitals or use of psychotropic medication. Information on hospital contacts was retrieved from the Danish Psychiatric Central Research Register (17) and the Danish National Patient Registry (18), which cover information on inpatient and outpatient contacts at psychiatric and somatic wards at Danish hospitals. Diagnoses included a wide range of mental and behavioral disorders. We excluded neurodevelopmental conditions (e.g. ADHD and autism spectrum disorders), given the potentially bidirectional interplay between these conditions and early adversity (19). Information on use of psychotropic medication was retrieved from the National Prescription Registry (20), which contains information on prescription medications dispensed at Danish pharmacies. Individuals with two or more prescriptions were classified as having mental health problems.

Lastly, information on criminal convictions was retrieved from the Danish System of Criminal Statistics (KRAF), which contains information on criminal charges and judicial decisions in Denmark. We included confirmed criminal convictions only. Codes included in each of the three outcomes are listed in the appendix (pp 8-10).

### Covariates

All analyses were adjusted for sex assigned at birth (female or male), year of birth, maternal age at birth (<20 years, 20–30 years, or >30 years), highest attained parental education (low [<10 years], middle [10–12 years], or high [>12 years]) and parental country of origin (Western [at least one parent from Europe, North America, Australia, or New Zealand] or non-Western [both parents from a non-Western country]). Further, all models included an interaction term between the childhood adversity group and a binary indicator denoting membership of the intervention subgroup (i.e., those individuals that were affected by the simulated intervention). This was included to capture potential heterogeneous effects of childhood adversity within this subgroup.

### Analytical framework

To explore the co-occurrence of poverty with other types of adversity, we first assessed how exposure to other adversities varied based on the number of years in childhood poverty.

We described rates of adversity within two key dimensions: loss or threat of loss (severe illness or death of sibling/parent) and family dynamics (e.g., parental drug/alcohol abuse, maternal separation or foster care placement). We further examined the prevalences of all three indicators of young adult social vulnerability by the number of years spent in childhood poverty.

The impact of the hypothetical 50% reduction in annual childhood poverty was explored using the following framework (as illustrated in Figure 1):

**Figure 1:**
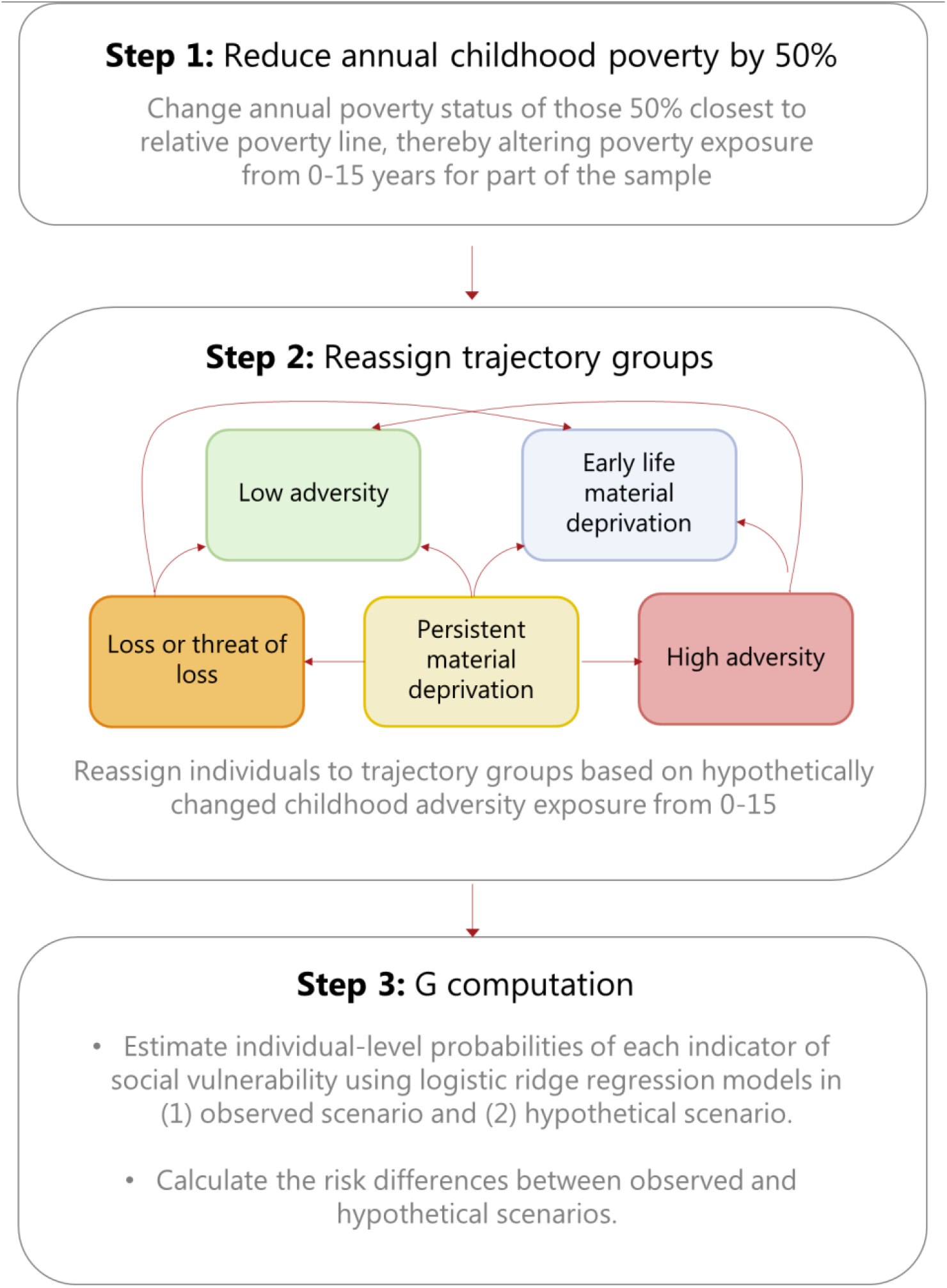
Overview of the methodological framework

#### Step 1

First, we hypothetically reduced the annual prevalence of childhood poverty by 50%. Each year, we reclassified the 50% of children living in poverty whose family incomes were closest to the ‘relative poverty line’ (i.e., 50% of the median income in the given year) from ‘poor’ to ‘not poor’. This approach was based on the assumption that those who are least poor are more likely to be lifted out of poverty by a poverty-reducing intervention.

#### Step 2

Next, we explored how such a hypothetical reduction in poverty would affect the distribution of individuals over patterns of childhood adversity in the population. We did this as childhood poverty is closely related to other types of adversities. Each child was re-assigned to one of the five childhood adversity groups based on their hypothetically changed exposure to poverty from ages 0-15, while their exposure within the loss or threat of loss and family dynamics dimensions was fixed to the observed level.

#### Step 3

The impact of the hypothetical poverty reduction on young adult social vulnerability was investigated using G-computation. We estimated individual-level probabilities of each of the three outcomes under both the observed and the simulated scenario. Then, we calculated risk differences for all three outcomes between the two scenarios, both in the entire population, and specifically among those affected by the hypothetical intervention. The 95% confidence intervals were estimated by bootstrapping for 1000 iterations. To mitigate multicollinearity issues, the models were fitted with ridge regression using the *glmnet* library in R (version 4.1-8). For each model, we selected the penalty parameter yielding the lowest mean cross-validated error using the *cv*.*glmnet* function.

## Results

Of the 607,308 individuals included in our study, 23% experienced poverty for at least one year from birth to age 15.

### Childhood poverty reduction and changes in childhood adversity

Children facing poverty were more likely to face additional adversities in childhood. Specifically, those spending more years in poverty were more likely to also experience loss or threat of loss in the family and adversities related to family dynamics (e.g., maternal separation, parental mental illness, foster care placement) (figure 2). Most strikingly, the rate of adversities within the family dynamics dimension was more than twenty times higher among those experiencing 15-16 years of poverty compared to those never experiencing poverty.

**Figure 2:**
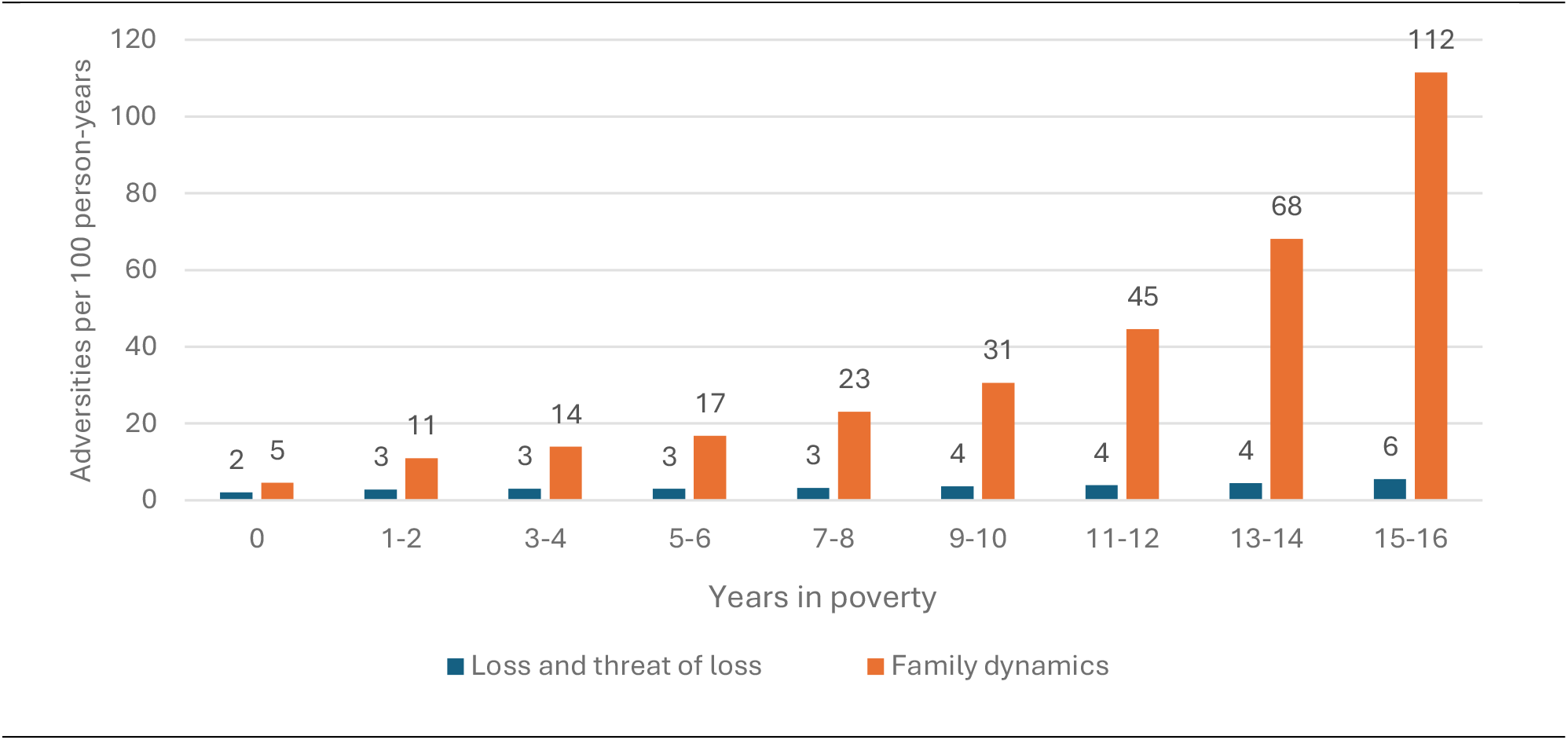
Rates of adversities across two dimensions (i.e., loss or threat of loss, and family dynamics) by years in poverty. *Individuals by years in poverty:* 0: n= 468,928 (77.7%) 1-2: n= 86,486 (14.2%), 3-4: n= 25,778 (4.2%), 5-6: n= 11,416 (1.9%), 7-8: n=6,170 (1.0%), 9-10: n= 3,625 (0.6%), 11-12: n= 2,201 (0.4%), 13-14: n= 1,526 (0.3%), 15-16: n= 1,178 (0.2%)

With this clustering of poverty with other adversities in mind, we explored how the distribution of individuals over the five childhood adversity groups changed following the hypothetical 50% reduction of childhood poverty, when fixing the level of exposure to other adversities to the observed level.

The hypothetical poverty reduction affected 17% of the population and resulted in a reduction in the proportion of individuals ever experiencing childhood poverty from about 23% to 13%. Following the hypothetical reduction in poverty, the proportion of individuals in the low adversity group increased from 52.6% to 55.3% (figure 3). The persistent material deprivation group decreased from 11.7 % to 9%. The early life material deprivation group, the loss and threat of loss group and the high adversity group all remained largely unchanged. A few individuals shifted from trajectories characterized mainly by material deprivation to those characterized by other forms of adversity, possibly reflecting the influence of other underlying adversities beyond material deprivation.

**Figure 3:**
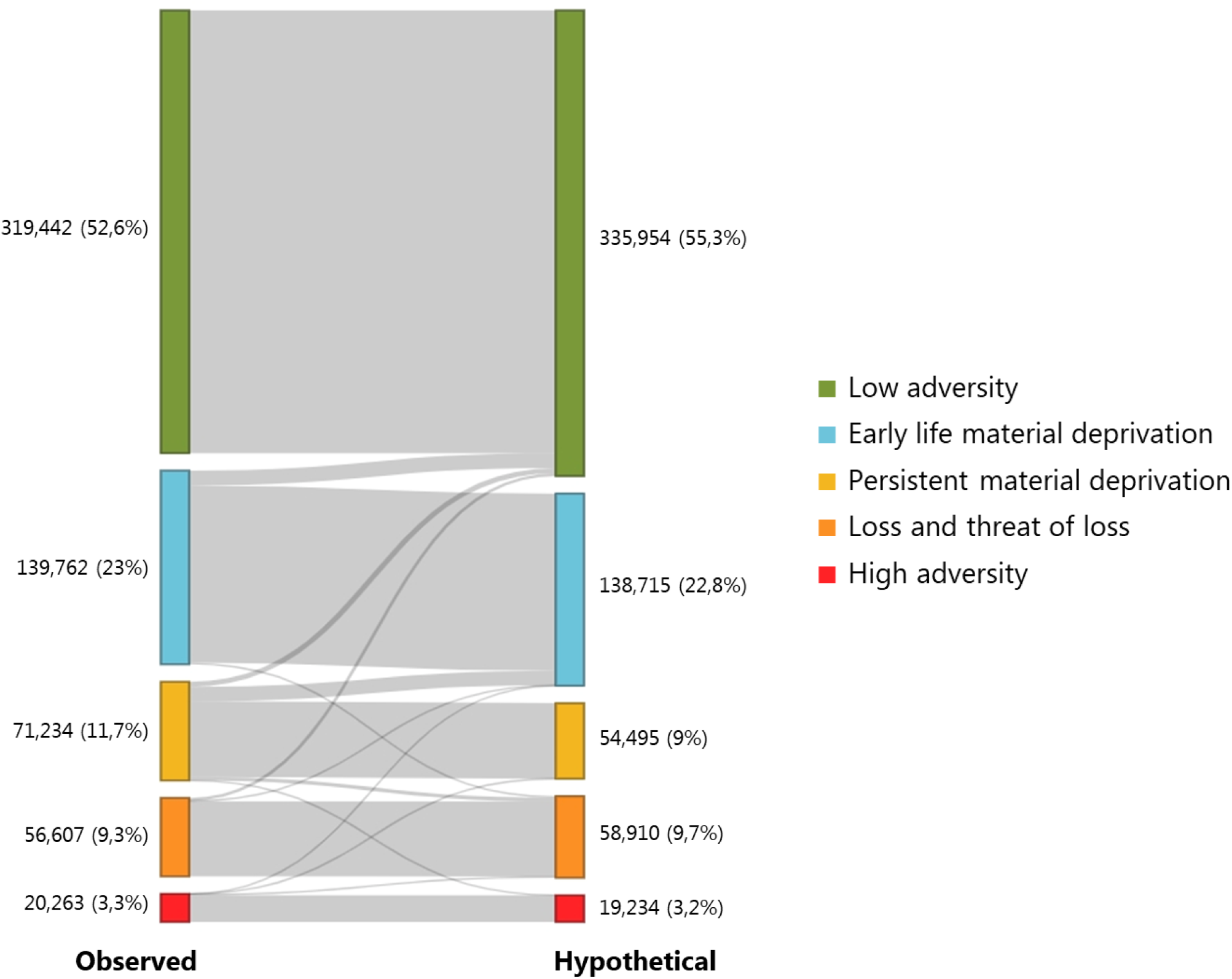
Sankey plot illustrating the distribution of individuals over five childhood adversity trajectory groups before and after the hypothetical poverty reduction. Group membership is based on the highest posterior probabilities estimated from the group-based trajectory model. The left side of the plot represents the distribution of individuals in the observed scenario, while the right side shows reassigned group memberships following the hypothetical intervention.

Among the individuals affected by the hypothetical intervention, 32% changed childhood adversity group, while the majority did not. Changes in adversity groups within this subgroup are illustrated in a Sankey plot in the appendix (p 11).

### Childhood poverty reduction and social vulnerability in young adulthood

All indicators of social vulnerability in young adulthood were more common among individuals who had been exposed to childhood poverty in the observed scenario (Figure 4). A gradient pattern was observed, with all three indicators being between two and four times more frequent among individuals exposed to childhood poverty for 15-16 years compared with those never exposed to poverty.

**Figure 4:**
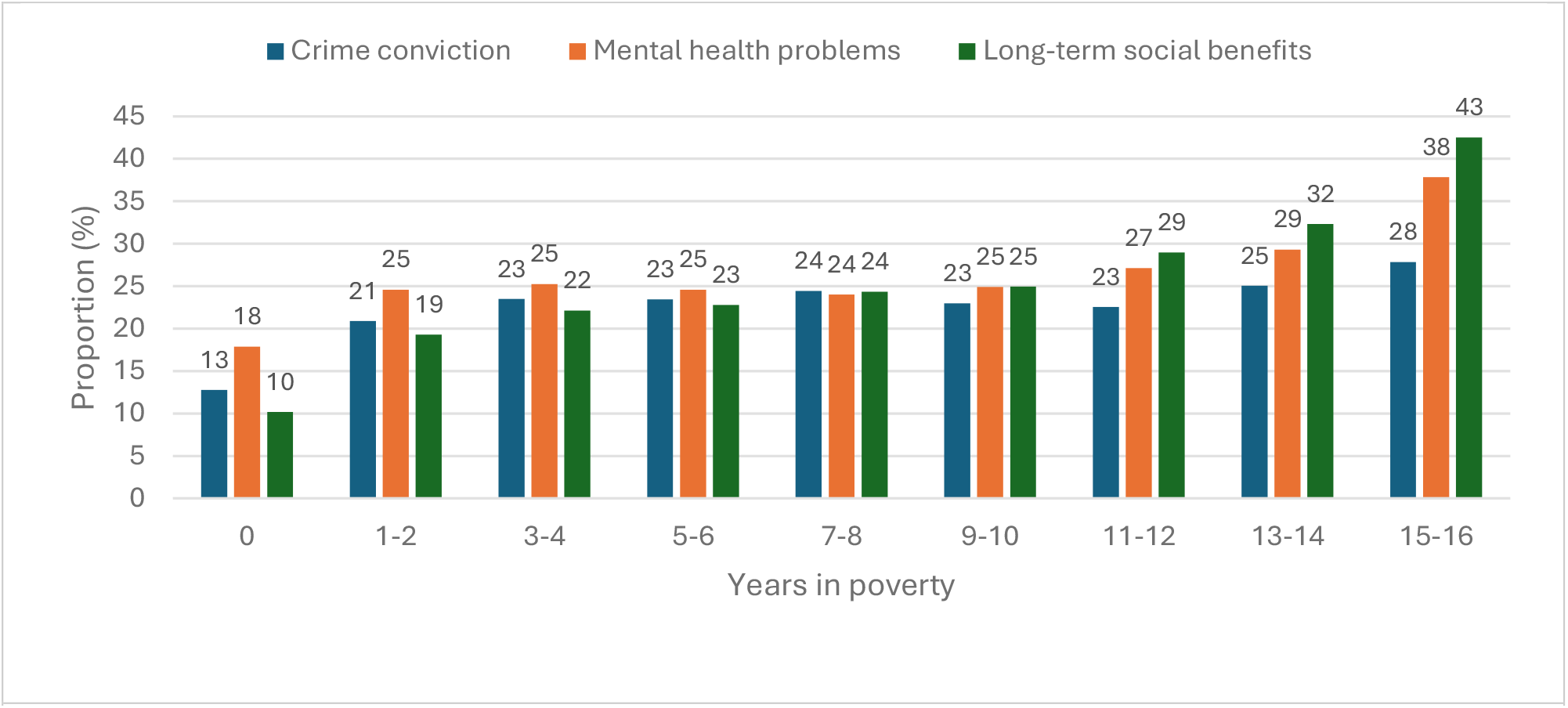
Proportion of study population with indicators of social vulnerability in young adulthood (i.e., crime conviction, mental health problems or long-term social benefit use) by years in childhood poverty

The hypothetical 50% reduction in childhood poverty was associated with a modest decrease in the occurrence of these three indicators of social vulnerability in the overall population (Figure 5). The largest reductions were observed for long-term social benefit use and criminal convictions, each decreasing by approximately 2 cases per 1,000 individuals (crime convictions: 1.9 fewer cases, 95% CI 1.8–2.0; long-term benefits: 2.1 fewer cases, 95% CI 2.0–2.2). Mental health problems decreased by about 1.2 cases per 1,000 individuals (95% CI 1.0–1.3). On a relative scale, these reductions corresponded to 1.7% fewer cases of long-term benefits, 1.3% fewer cases of crime convictions, and 0.6% fewer cases of mental health problems.

**Figure 5:**
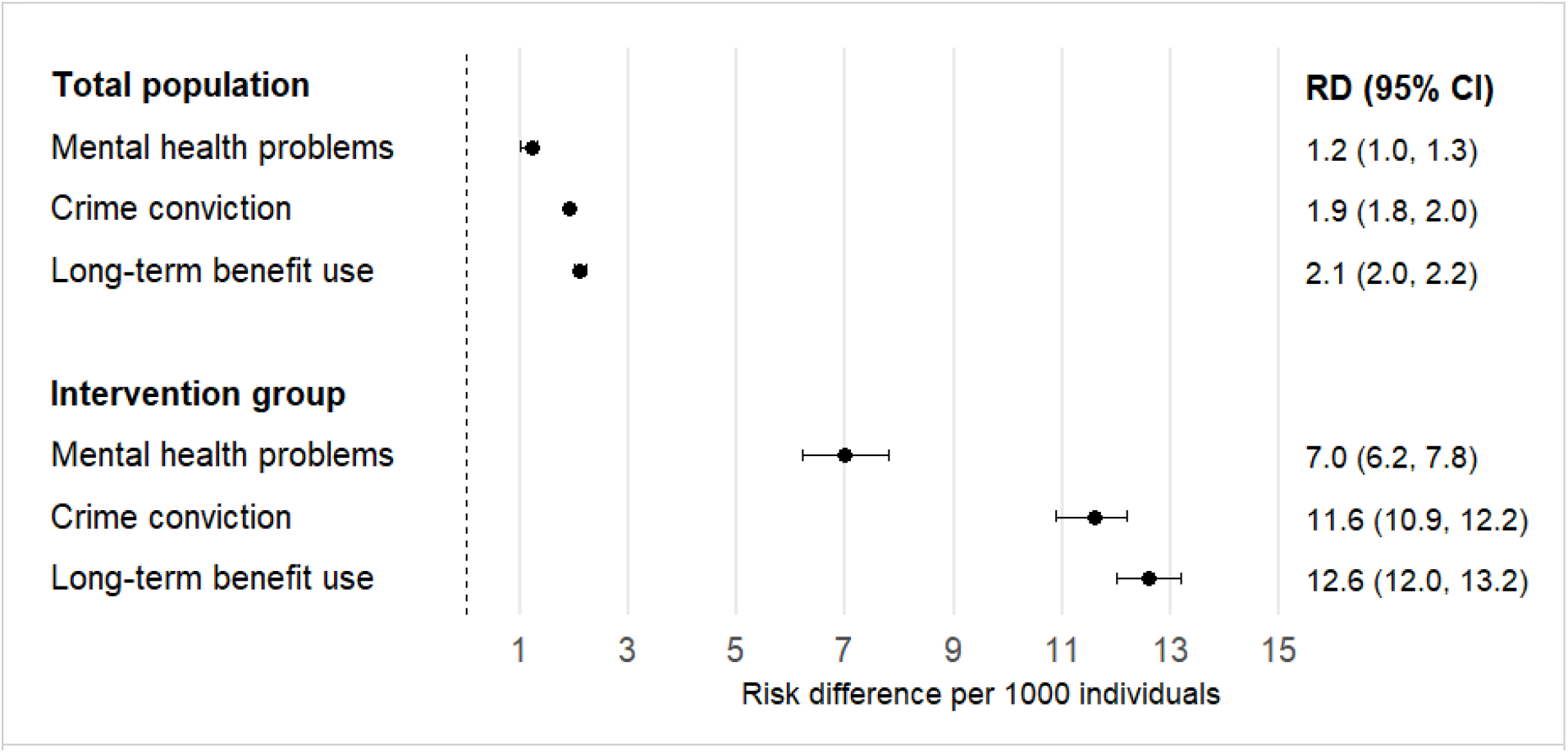
Risk differences per 1000 individuals in the total population and in the subgroup being affected by the hypothetical intervention.

Among the 17% of the population who was affected by the hypothetical reduction, we identified larger reductions in social vulnerability with 7.0 (CI95%: 6.2;7.8) fewer cases of mental health problems, 11.6 (CI95%: 10.9;12.2) fewer cases of crime conviction and 12.6 (CI95%: 12.0;13.2) fewer cases of long-term benefits per 1,000 individuals. These reductions corresponded to 3% fewer individuals with mental health problems, 5.4% fewer convicted of a crime and 6.4% fewer with long-term benefit use. In a sensitivity analysis in which individuals were weighted by their posterior probabilities of group membership, we observed similar reductions in both the total population and the intervention group (results not shown). Average latent class posterior probabilities of group assignment under observed and hypothetical scenarios are presented in the appendix (p 12).

## Discussion

Aligning with the SDG goal of reducing childhood poverty, this nationwide study explores the impact of hypothetically reducing annual childhood poverty by 50%. We demonstrate that children experiencing more years of poverty were also more likely to experience higher rates of other adversities during childhood and to experience social vulnerability in young adulthood. Hypothetically reducing childhood poverty by 50% led to changes in the distribution of individuals over trajectories of childhood adversity, with most shifts occurring from a trajectory characterized by persistent material deprivation to one characterized by early material deprivation, and from early material deprivation to low adversity. However, in the group affected by the hypothetical intervention, most children did not change trajectory groups, emphasizing that the patterns of childhood adversity are not determined by poverty alone.

In accordance with our hypothesis, we find that hypothetically reducing childhood poverty by 50% was associated with lower risk of social vulnerability in young adulthood. In the intervention group, the association corresponded to 7 fewer individuals with mental health problems, 12 fewer individuals with a crime conviction, and 13 fewer individuals with long-term social benefit use per 1000 persons.

Our findings align with studies demonstrating that children in poverty are at higher risk of mental health disorders (4), becoming involved in crime (3), and being marginalized in the labor market while transitioning into adulthood (21). The scientific literature has long sought to determine whether such inequalities reflect a causal relationship. A range of experimental and quasi-experimental studies have suggested that childhood poverty affects well-being and development during childhood (10). However, fewer studies have focused specifically on the long-term consequences reaching into adulthood (11). Some quasi-experimental studies have suggested that alleviating childhood poverty is associated with reduced social vulnerability in young adulthood, including lower rates of mental health disorders, fewer criminal convictions and lower risk of labor market marginalization (21–23) while others found no such effect (24). Yet, given that such studies are often based on narrowly defined or selective study populations, their findings may be less generalizable to broader contexts. The current study adds to the previous literature by providing population-wide estimates on the hypothetical impact of reducing childhood poverty in the context of a welfare state, while addressing the cooccurrence of other adversities, which may also contribute to social vulnerability in young adulthood.

The current study emphasizes the strong accumulation of various forms of adversity among children facing poverty. This finding is consistent with the cumulative disadvantage theory which posits that disadvantages tend to cluster within certain social groups, compounding over time and leading to increasingly adverse outcomes (25). From this perspective, reducing childhood poverty may mitigate the risk of additional disadvantages during childhood and later in the life course.

Material deprivation has been suggested to influence the developing child by limiting access to basic needs fundamental for healthy development and thriving, including quality housing, healthy foods, and prescription medicine. Furthermore, childhood poverty has been associated with family stress, which may influence parenting style and the overall home environment (26,27). While we controlled for the level of other adversities, such as parental mental health, we were unable to fully capture potential stress-related mechanisms linking childhood poverty to social vulnerability later in life. Additionally, experiences and values in families facing poverty may shape children’s attitudes toward, for example, employment and legal norms, potentially influencing their likelihood of labor market participation and involvement in crime as young adults (3).

Our findings suggest that even in a welfare state with a generous social security system such as the Danish, reducing childhood poverty may have long-term effects on social vulnerability. While these estimates cannot be directly translated into other contexts, they may provide a ‘lower bound scenario’ and the estimated benefits are likely to be more pronounced in countries lacking social security, where children in poverty face greater material hardship and limited access to healthcare (28).

## Strengths & limitations

The current study can be perceived as a globally unique ‘data laboratory’ where an innovative methodological framework is combined with empirical data from a large, unselected study population, allowing us to ask policy-relevant *what if* questions about poverty reductions and assess their benefits. The methodological framework integrates group-based multi-trajectory modelling and G-computation, which allowed for a thorough investigation into how the distribution over patterns of childhood adversity might change following a hypothetical reduction in childhood poverty. Most epidemiological studies report ‘exposure effects’ which compare individual risk with and without a given exposure. Such estimates are useful for evaluating the effects of lifestyle or clinical treatments on health. However, they are less relevant for assessing the impact of population-level interventions, where it is often unrealistic for everyone or no one to be exposed (29). By applying the parametric G-formula, we were able to estimate the potential effect of a population intervention, thereby providing policy-relevant insights into the effect of reducing childhood poverty.

Further, the high-quality data enabled the development of a comprehensive measure of childhood adversity drawing on detailed and temporally ordered information on 12 adversities across three central dimensions from birth to age 15. A limitation of this measure is that information on abuse, neglect, and domestic violence was not available through the Danish registries. Yet, due to the accumulation and social patterning of adversity, we believe we have captured the overall patterns of childhood adversity.

In acknowledging that childhood poverty is highly intertwined with other adversities, we fixed the levels of other types of adversities such as parental mental health and unemployment to their observed levels. By doing so, we were unable to fully capture the potential effects that a reduction in childhood poverty might have on preventing other adversities. Previous studies have suggested that poverty reduction may have a positive impact on a range of factors critical for healthy development during childhood—for example, by improving parental mental health (10,30). From this perspective, our study may provide conservative estimates of the effects of poverty reductions on social vulnerability in young adulthood, especially for individuals in childhood adversity groups characterized by other adversities beyond material deprivation (i.e., the loss and threat of loss and high adversity group).

Our results may be affected by unmeasured confounding from differences in values and beliefs within families. By adjusting for parental education, we have likely captured some differences in family attitudes toward education, employment, and beliefs about criminal behavior. However, we have likely not fully addressed this issue.

## Perspectives & conclusion

This study demonstrates that, even within a welfare state, reducing childhood poverty may lower young adult social vulnerability. As we likely provided conservative estimates of the long-term benefits of reducing childhood poverty on social vulnerability in young adulthood, the actual benefits of reducing childhood poverty may be greater, particularly in contexts with less extensive social security systems. Potential interventions to reduce childhood poverty could include universal policies to increase child benefits or unemployment transfers to families, as well as measures to reduce the costs of basic needs such as housing.

As childhood poverty reduction is unlikely to eliminate exposure to other adversities, strategies to reduce poverty should be integrated with other initiatives addressing the multifaceted adversities associated with poverty. This includes multi-sector policies that provide support for families facing adversity, such as mental or somatic health issues, loss in the family or parental separation. Addressing such interconnected factors is crucial to breaking the cycles of poverty and other adversities and reducing young adult social vulnerability.

## Data Availability

The data material contains personally identifiable and sensitive information. According to the Act on Processing of Personal Data, such data cannot be made publicly available.

## Notes

### Competing Interest Statement

The authors have declared no competing interest.

### Author Declarations

Every Danish citizen has a unique personal identification number, which allows for linkage between various administrative and health registers. This unique opportunity for data linkage will be utilized to build the DANLIFE-2 cohort. The cohort include everyone born in Denmark since 1980, including potentially vulnerable groups like children or socially marginalized groups. The health registers also include sensitive information about health histories. To prevent misuse of any of these data, the Equal Health project follow strict national data security rules and legislation to protect personal data. Access to the administrative and research registers in Denmark is obtained through Statistics Denmark (http://www.danmarksstatistik.dk/en/TilSalg/Forskningsservice.aspx), which provides information from administrative registers, and Sundhedsdatastyrelsen (https://sundhedsdatastyrelsen.dk/da/forskerservice), which provides access to register-based health data. Only Danish research institutions can be granted authorization to access these data, and the data can only be used for research purposes. All data is handled according to GDPR guidelines. We have a formalized training system in GDPR and data safety, which is mandatory for all researchers working with data at the University of Copenhagen. Data access is provided through remote access to a secure server placed at Statistics Denmark. The remote access is given via a protected and encrypted internet access. The research server is separated from the production network and the data is only accessible in de-identified form. The DANLIFE study is approved by the Danish Data Protection Agency through the records of research projects (involving personal data) at the Faculty of Health and Medical Sciences, University of Copenhagen (record no 514-0641/21-3000). The Danish Data Protection Agency (datatilsynet.dk/english) ensures compliance with national and EU legislation. I will seek renewed approval for the planned extension into DANLIFE-2 for the Equal Health project. Register-based studies do not require ethical approval by the Danish National Committee on Health Research Ethics according to Danish Law (Section 14, part 2) because 'Notification of questionnaire surveys and medical database research projects to the system of research ethics committee system is only required if the project involves human biological material' (https://en.nvk.dk/rules-and-guidelines/act-on-research-ethics-review-of-health-research-projects).

