## Supplementary Materials for "Impact of reducing childhood poverty on social vulnerability in young adulthood: a simulation study based on nationwide life-course data"

### SUPPLEMENTARY MATERIAL

#### Overview

**Supplementary Table 3.** Overview of included mental health diagnoses (from the Psychiatric Central Research Register and DNPR) and prescription medications (from the National Prescription Registry). ... 9

**Supplementary Table 4.** Overview of convicted crimes included in the study from the KRAF register . 10

**Supplementary Figure 3.** Sankey plot, intervention subgroup ........ 11

**Supplementary Table 5.** Average latent posterior probabilities of group assignment from Group-Based Multi-Trajectory Modeling ........ 12

**Supplementary Figure 1.** Flow chart

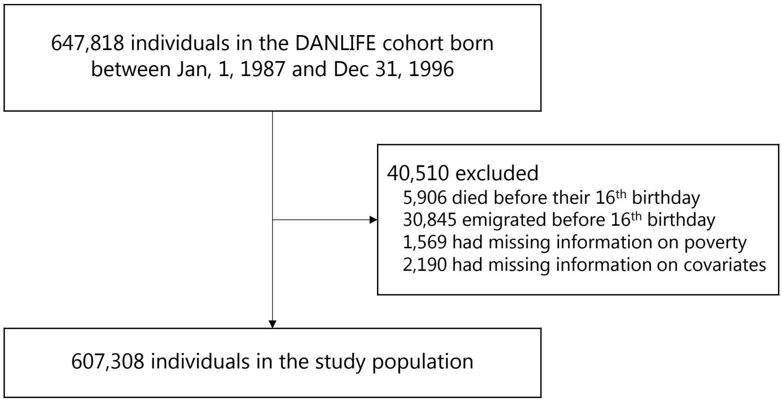

**Supplementary Table 1.** Dimensions and definitions of childhood adversities in the five trajectory groups of adversity identified by Rod et al. (1). Details described in Bengtsson et al. (2). The authors on Rod et al. (1) included experts in stress, child health, and child psychology and this panel of experts decided on the three predefined dimensions of childhood adversity.

| | Adversity | Definition | Registers |
|---|---|---|---|
| Material deprivatio | **Family poverty** | Family income below 50% of the median national family income in a given year | The Income Statistics Register (3) |
| | **Parental long-term unemployment** | A parent being unemployed for at least 12 months | The Integrated Database for Labour Market Research (4) |
| Loss or threat of loss | **Death of a parent** | Death of a parent | The Danish Civil Registration System (5) |
| | **Death of a sibling** | Death of a sibling | The Danish Civil Registration System (5) |
| | **Parental somatic illness** | A parent being diagnosed with one of the diseases included in the Charlson comorbidity index | The Danish National Patient Register (6) |
| | **Sibling somatic illness** | A sibling being diagnosed with one of the seven somatic illnesses most commonly related to mortality in children aged 0-18 years in Denmark: malignant neoplasm; congenital anomalies of the heart and circulatory system; congenital anomalies of the nervous system; cerebral palsy; epilepsy; cardiomyopathy; congenital disorders of lipid metabolism | The Danish National Patient Register (6) |
| Family dynamics | **Foster care** | Being placed in out-of-home care | The Register of Support for Children and Adolescents (7) |
| | **Parental psychiatric illness** | A parent being admitted for at least 1 day to a psychiatric hospital or ward with a primary diagnosis related to psychiatric illness (excluding primary diagnoses related to alcohol and drug abuse) | The Danish Psychiatric Central Research Register (8); The Danish National Patient Register (6) |
| | **Sibling psychiatric illness** | A sibling being admitted for at least 1 day to a psychiatric hospital or ward with a primary diagnosis related to psychiatric illness | The Danish Psychiatric Central Research Register (8); The Danish National Patient Register (6) |
| | **Parental alcohol abuse** | A parent being diagnosed with a disease related to alcohol abuse or buying a prescribed drug used in treatment of alcohol dependence | The Danish Psychiatric Central Research Register (8); The Danish National Patient Register (6); The Danish National Prescription Registry (9) |
| | **Parental drug abuse** | A parent being diagnosed with a disease related to drug abuse or buying a prescribed drug used in treatment of drug dependence | The Danish Psychiatric Central Research Register (8); The Danish National Patient Register (6); The Danish National Prescription Registry (9) |
| | **Maternal separation** | The mother no longer sharing address with a partner | The Danish Civil Registration System (5) |

**Note:** This table is replicated from the supplementary material of Rod NH, Bengtsson J, Elsenburg LK, Taylor-Robinson D, Rieckmann A. Hospitalisation patterns among children exposed to childhood adversity: a population-based cohort study of half a million children. Lancet Public Health 2021; published online Sept 29. http://dx.doi.org/10.1016/S2468-2667(21)00158-4.

References to Supplementary Table 1

**Supplementary Figure 2.** Annual rates of childhood adversity in the five trajectory groups from age 0-15 across three predefined dimensions; material deprivation (upper panels), loss or threat of loss (middle panels) and family dysfunction (lower panels). The annual rates are presented as events per person-year. The columns represent the five trajectory groups: Low Adversity (green), Early Life Material Deprivation (blue), Persistent Material Deprivation (yellow), Loss or Threat of Loss (orange) and High Adversity (red). The percentages refer to the percentage of children belonging to each trajectory group in the sample, e.g., 3% of the children belong to the high adversity group. See Supplementary Table 1 for a detailed description of the adversities included in each of the three dimensions.

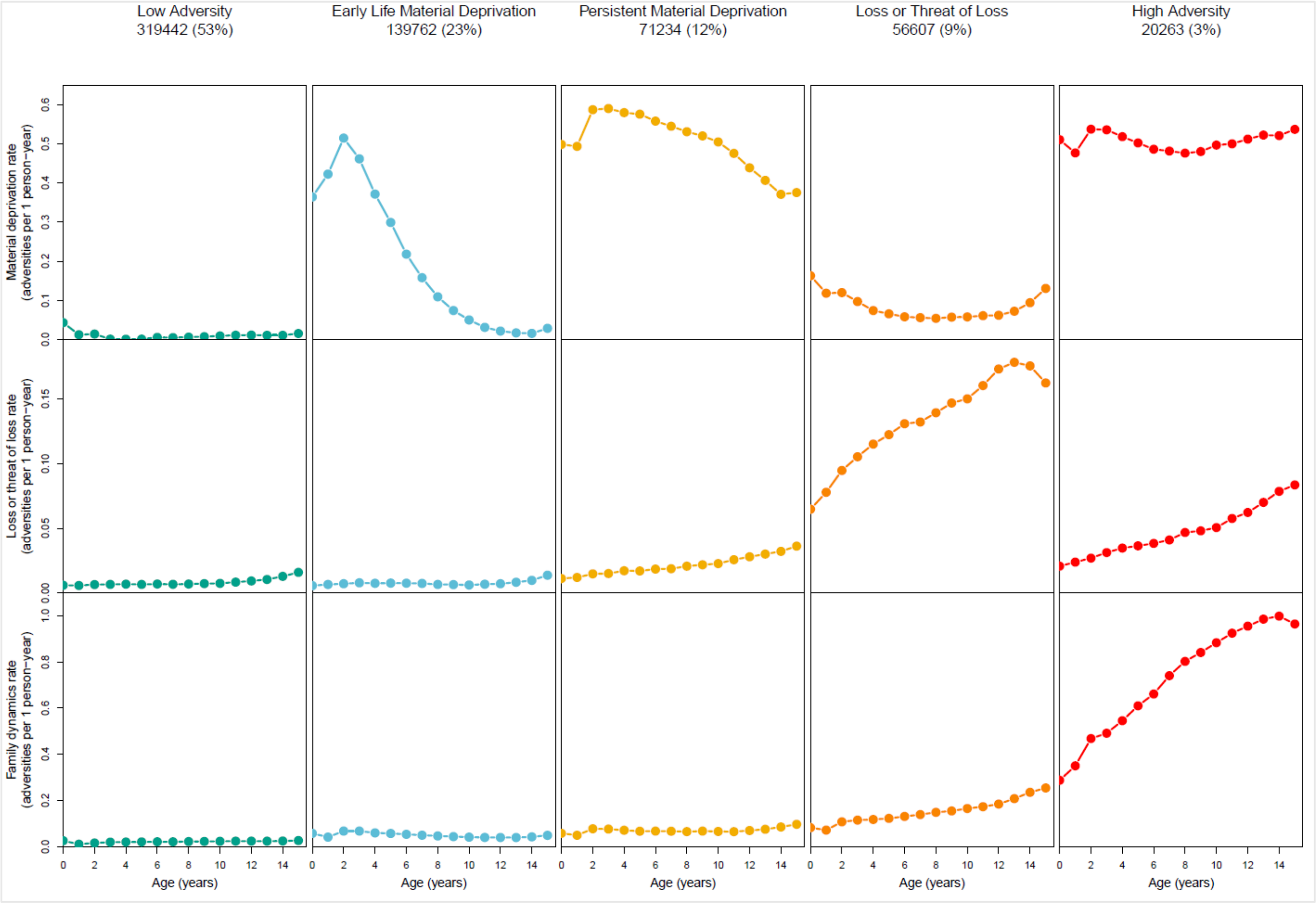

**Supplementary Methods.** Group-based multi-trajectory model

A group-based multi-trajectory modeling was applied following the approach in Nagin, D. S., Jones, B. L., Passos, V. L., & Tremblay, R. E. (2018). Group-based multi-trajectory modeling. Statistical methods in medical research, 27(7), 2015- 2023. We aimed to determine the most common trajectory groups of childhood adversity based on adversities in three predefined dimensions: material deprivation, loss or threat of loss, and family dynamics. For each child, we summed the number of annual childhood adversity experiences per dimension as illustrated in the hypothetical example below depicting one imaginary child:

*Hypothetical example of one study participant*

| ***Dimension*** | ***Event type*** | ***Age 0-1*** | ***Age 1-2*** | ***(…)*** | ***Age 15-16*** |
|---|---|---|---|---|---|
| Material deprivation | Family poverty | 1 | 0 | | 1 |
| | Long-term unemployment | 0 | 2 | | 1 |
| **Sum vector for dimension** | | **1** | **2** | | **2** |
| Loss or threat of loss | Death of a parent | 1 | 0 | | 0 |
| | Death of a sibling | 0 | 1 | | 0 |
| | Parental somatic illness | 0 | 2 | | 0 |
| | Sibling somatic illness | 1 | 0 | | 0 |
| **Sum vector for dimension 2** | | **2** | **3** | | **0** |
| Family dynamics | Foster care | 2 | 0 | | 0 |
| | Parental psychiatric illness | 0 | 0 | | 0 |
| | Sibling psychiatric illness | 0 | 0 | | 0 |
| | Parental alcohol abuse | 0 | 0 | | 0 |
| | Parental drug abuse | 0 | 0 | | 0 |
| | Maternal seperation | 0 | 0 | | 0 |
| **Sum vector for dimension 3** | | **2** | **0** | | **0** |

As outlined above, each adversity experience only counts in the year in which it occurs, but the same type of adversity can occur multiple times during childhood. As an example, see the cell in column “Age 1-2” row “Parental somatic illness”. Here, having a mother with cancer will count towards the ‘loss or threat of loss’ dimension only in the years in which the mother has a cancer diagnosis in the hospital register. Additionally, having two parents with cancer within the same year will result in a count of two in the giving year. Thus, this imaginary participant has two parents with hospital contacts for severe illnesses during his/her second year of live.

We used the package TRAJ for Stata to fit group-based multi-trajectory models using zero-inflated Poisson regressions and modeled the trajectories with a cubic function of age. The model yielded a probability for each individual of being in each trajectory group.

Due to computational issues with large data, we employed a 2-stage approach. First, we fitted a model based on a random sample of 50,000 individuals. Second, we extrapolated the estimated probabilities of being in each trajectory group onto the full cohort. To assess the consistency of estimating the model on a sub-sample, we re-ran the procedure on 5 random samples of 50,000 individuals selected from the total study population, which returned five almost identical estimated models.

**Note:** The supplementary methods are replicated from the supplementary material of Rod NH, Bengtsson J, Elsenburg LK, Taylor-Robinson D, Rieckmann A. Hospitalisation patterns among children exposed to childhood adversity: a population-based cohort study of half a million children. Lancet Public Health 2021; published online Sept 29. http://dx.doi.org/10.1016/S2468-2667(21)00158-4.

**Supplementary Table 2.** Overview of social welfare codes included in the study (DREAM register)**.**

| Dream code | Benefit type (Danish name of benefit) |
|---|---|
| 111, 213-219, 299 | Unemployment benefit (‘Dagpenge’) |
| 130-139, 730-739 | Social assistance (‘Kontanthjælp’) |
| 151 | Special educational support (‘Særlig uddannelsesydelse’) |
| 152 | Labour market benefit (‘Arbejdsmarkedsydelse’) |
| 153 | Cash benefit (‘Kontantydelse’) |
| 700-709, 160-169 | Integration benefit (‘Integrationsydelse’) |
| 740-748 | Unemployment support (‘Ledighedsydelse’) |
| 750-758 | Pre-rehabilitation benefit (‘For revalidering’) |
| 760-768 | Rehabilitation benefit (‘Revalidering’) |
| 783 | Early retirement pension (‘Førtidspension’) |
| 784 | Early retirement pension/Social assistance (‘Førtidspension/kontanthjælp’) |
| 810-818 | Resource course benefit (‘Ressourceforløbsydelse’) |
| 870-878 | Job assessment (‘Jobafklaring’) |
| 890, 893-899 | Sickness benefit (‘Sygedagpenge’) |

*Note: The names of some of the benefits were translated for the purpose of this study*

**Supplementary Table 3.** Overview of included mental health diagnoses (from the Psychiatric Central Research Register and DNPR) and prescription medications (from the National Prescription Registry).

| ICD-10 codes | Diagnoses |
|---|---|
| F10-F19.9 | Mental and behavioural disorders due to psychoactive substance use |
| F20-F29 | Schizophrenia, Schizotypal, delusional, and other non-mood psychotic disorders |
| F30-F31.9, F32-F34.0 and F38-F39 | Mood [Affective] disorders |
| F40-F48.9 | Anxiety, dissociative, stress-related, somatoform and other non-psychotic mental disorders |
| F50-F51.9 | Behavioral syndromes associated with physiological disturbances and physical factors |
| F60-F61 | Disorders of adult personality and behavior |
| **ATC code** | **Medication** |
| N05A | Antipsychotics |
| N0BA | Antidepressants |
| N0BA | Anxiolytics (except for N0BA: Benzodiazepine derivatives) |
| N05AN | Lithium |
| N07BB | Alcohol dependence medication |
| N07BC | Opiod dependence medication |

**Supplementary Table 4.** Overview of convicted crimes included in the study from the KRAF register

| 2-digit codes (variable AFG_GER7) | Type of crimes (Danish name of the crime) |
|---|---|
| 10 | Undisclosed criminal law (Uoplyst straffelov) |
| 11 | Sexual crimes (Seksualforbrydelser) |
| 12 | Violent crimes (Voldsforbrydelser) |
| 13 | Property crimes (Ejendomsforbrydelser) |
| 14 | Other crimes (Andre forbrydelser Færdselslov) |
| 32 | Act on euphoric substances (Lov om euforiserende stoffer) |
| 34 | Weapons Act (Våbenloven) |
| 36 | Tax and levy laws (Skatte- og afgiftslove) |
| 38 | Other special laws (Særlove i øvrigt) |

*Note: The names of some of the benefits were translated for the purpose of this study*

**Supplementary Figure 3.** Sankey plot, intervention subgroup

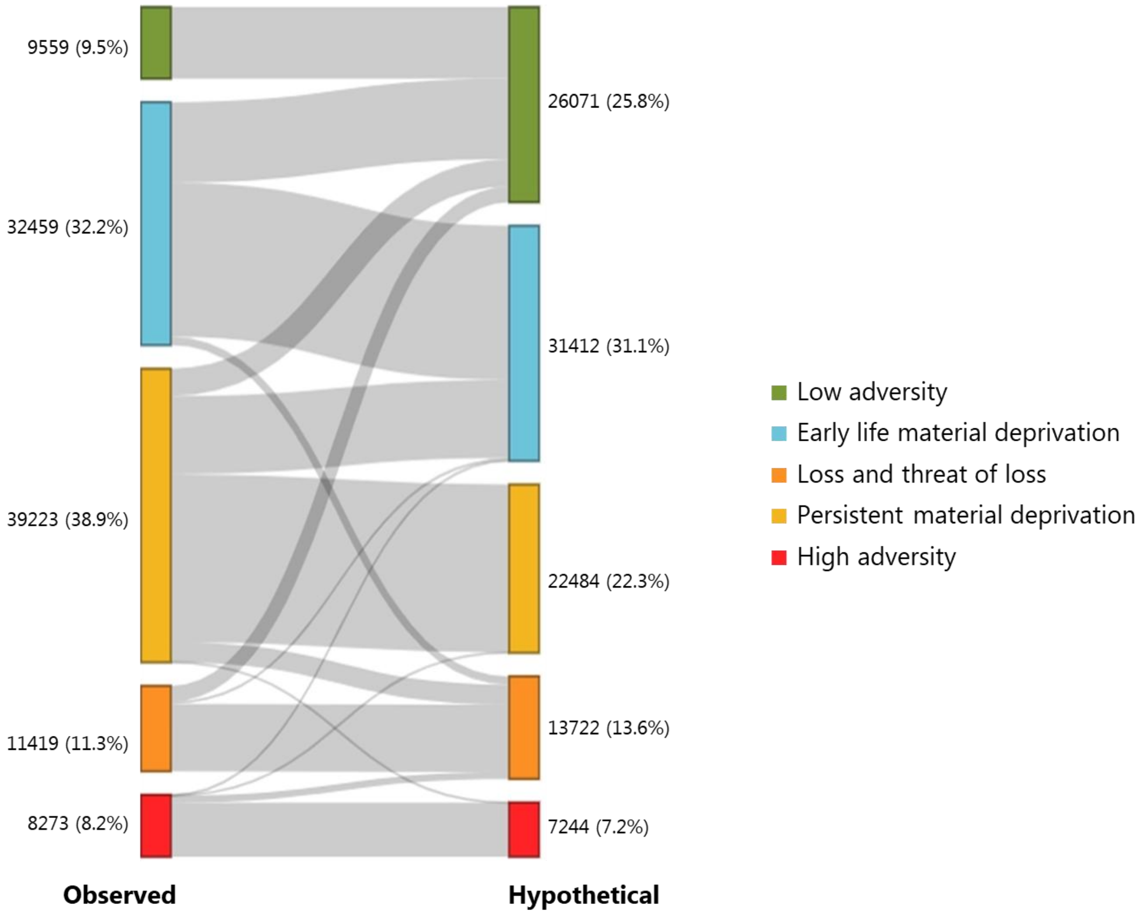

**Note:** Sankey plot illustrating changes in the distribution of individuals across five trajectory groups before and after the hypothetical poverty reduction. Group membership is based on the higest posterior probabilities estimated from the group-based trajectory model. The left side of the plot represents the distribution of individuals in the observed scenario, while the right side shows reassigned group memberships following the hypothetical intervention.

**Supplementary Table 5.** Average latent posterior probabilities of group assignment from Group-Based Multi-Trajectory Modeling under observed and hypothetical scenarios, shown for the total population and for individuals affected by the hypothetical intervention.

| Total population | | | | | | |
|---|---|---|---|---|---|---|
| **Trajectory group** | **Observed scenario** | | | **Hypothetical scenario** | | |
| | *n* | *Mean* | *SD* | *n* | *Mean* | *SD* |
| Low adversity | 319442 | 0,918004312 | 0,120465389 | 335954 | 0,918041 | 0,119892 |
| Early life material deprivation | 139762 | 0,833504007 | 0,165825028 | 138715 | 0,834933 | 0,165935 |
| Persistent deprivation | 71234 | 0,873202199 | 0,164154929 | 54495 | 0,856499 | 0,168661 |
| Loss or threat of loss | 56607 | 0,828242843 | 0,177500964 | 58910 | 0,832681 | 0,175248 |
| High adversity | 20263 | 0,934660358 | 0,130361868 | 19234 | 0,935641 | 0,12952 |

| Intervention group | | | | | | |
|---|---|---|---|---|---|---|
| **Trajectory group** | **Observed scenario** | | | **Hypothetical scenario** | | |
| | *n* | *Mean* | *SD* | *n* | *Mean* | *SD* |
| Low adversity | 9559 | 0,733865713 | 0,163666333 | 26071 | 0,850956 | 0,158657 |
| Early life material deprivation | 32459 | 0,804600456 | 0,167805784 | 31412 | 0,809948 | 0,169289 |
| Persistent deprivation | 39223 | 0,893099119 | 0,155468954 | 22484 | 0,867428 | 0,164537 |
| Loss or threat of loss | 11419 | 0,792898285 | 0,190236182 | 13722 | 0,817883 | 0,181682 |
| High adversity | 8273 | 0,933163776 | 0,132221544 | 7244 | 0,935556 | 0,130289 |
